# Gaps in the phenotype descriptions of ultra-rare genetic conditions: review and multicenter consensus reporting guidelines

**DOI:** 10.1101/2023.09.13.23295418

**Authors:** Ali AlMail, Ahmed Jamjoom, Amy Pan, Min Yi Feng, Vann Chau, Alissa D’Gama, Katherine Howell, Nicole S.Y. Liang, Amy McTague, Annapurna Poduri, Kimberly Wiltrout, IPCHiP Executive Committee, Anne S. Bassett, John Christodoulou, Lucie Dupuis, Peter Gill, Tess Levy, Paige Siper, Zornitza Stark, Jacob A.S. Vorstman, Catherine Diskin, Natalie Jewitt, Danielle Baribeau, Gregory Costain

**Affiliations:** Temerty Faculty of Medicine, University of Toronto, Toronto, Canada; Program in Genetics & Genome Biology, SickKids Research Institute, Toronto, Canada; Department of Paediatrics, University of Toronto, Toronto, Canada; Department of Molecular Genetics, University of Toronto, Toronto, Canada; Division of Neurology, Hospital for Sick Children, Toronto, Canada; Department of Neurology, Boston Children’s Hospital, Boston, MA, USA; Division of Newborn Medicine, Department of Pediatrics, Boston Children’s Hospital, Boston, MA, USA; Department of Pediatrics, Harvard Medical School, Boston, MA, USA; Department of Neurology, Royal Children’s Hospital, Melbourne, Australia; Murdoch Children’s Research Institute, Melbourne, Australia; Division of Clinical and Metabolic Genetics, Hospital for Sick Children, Toronto, Canada; Department of Neurology, Great Ormond Street Hospital, London, UK; Developmental Neurosciences, UCL Great Ormond Street Institute of Child Health, London, UK; Department of Neurology, Harvard Medical School, Boston, MA, USA; Department of Psychiatry, University of Toronto, Toronto, Canada; Division of Psychiatry, Ichan School of Medicine at Mount Sinai, New York City, New York, United States of America; Department of Paediatrics, University of Melbourne, Melbourne, Australia; Victorian Clinical Genetics Service, Melbourne, Australia; Department of Psychiatry, Hospital for Sick Children, Toronto, Canada; Autism Research Centre, Holland Bloorview Kids Rehabilitation Hospital, Toronto, Canada

**Keywords:** Phenotype, Ultra-rare genetic conditions, Genome sequencing, Natural history, Mendelian genetic diseases

## Abstract

**Background:** Genome-wide sequencing and genetic matchmaker services are propelling a new era of genotype-first ascertainment of novel genetic conditions. The degree to which reported phenotype data in discovery-focused studies address informational priorities for clinicians and families is unclear.

**Methods:** We identified reports published from 2017-2021 in ten genetics journals of novel Mendelian disorders ascertained genotype-first. We adjudicated the quality and detail of the phenotype data via 46 questions pertaining to six priority domains: (I) Development, cognition, and mental health; (II) Feeding and growth; (III) Medication use and treatment history; (IV) Pain, sleep, and quality of life; (V) Adulthood; and (VI) Epilepsy. For a subset of articles, all subsequent published follow-up case descriptions were identified and assessed in a similar manner. A modified Delphi approach was used to develop consensus reporting guidelines, with input from content experts across four countries.

**Results:** In total, 200 of 3243 screened publications met inclusion criteria. Relevant phenotypic details across each of the six domains were rated superficial or deficient in >87% of papers. For example, less than 10% of publications provided details regarding neuropsychiatric diagnoses and “behavioural issues”, or about the type/nature of feeding problems. Follow-up reports (n=95) rarely addressed the limitations of the original reports. Reporting guidelines were developed for each domain.

**Conclusion:** Phenotype information relevant to clinical management, genetic counseling, and the stated priorities of patients and families is lacking for many newly described genetic diseases. Use of the proposed guidelines could improve phenotype reporting in the genomic era.

## INTRODUCTION

Genome-wide sequencing and genetic matchmaker services have created a new paradigm for Mendelian disorder delineation.^1-3^ Compared to prior decades, when syndrome identification was predominantly phenotype driven, there is now an increasing focus on genotype-first ascertainment and on generating functional evidence or usage of non-human model systems to support the disease-variant/gene association. There are no well-defined nor broadly accepted minimum standards for phenotype descriptions of putative novel disorders with multisystem manifestations and/or a neurodevelopmental component. Variability in the consistency of phenotyping and describing of findings is problematic. This variability can be exacerbated by individual sites each contributing only a single patient to an international case-series study, or the extraction of phenotype data from laboratory test requisitions.

After a first description of a novel Mendelian disorder is published, patients soon thereafter begin to be diagnosed via clinical genome-wide sequencing.^4-6^ These individually ultra-rare^7^ conditions are collectively important contributors to the burden of genetic disease in the population.^8^ The typical benefits of a molecular genetic diagnosis^7,9^ are attenuated when there is limited information available to inform genotype-phenotype correlation, natural history, prognostication, and anticipatory care. A key consideration in the assessment of ultra-rare conditions for potential “precision therapy” development is the degree to which the patient’s clinical trajectory can be anticipated.^10,11^ Families who are amongst the first to receive a diagnosis of an ultra-rare genetic disorder have endorsed frustration with the perceived lack of information and support.^12,13^ Similarly, clinicians face the same informational barrier, which impacts their abilities to care for and counsel patients and their families.^14^

Published expert opinions, survey data, reviews, and data from patient and family focus groups highlight key informational areas germane to the natural history of ultra-rare genetic diseases.^14-24^ We assessed the breadth and depth of phenotype reporting in contemporary descriptions of novel Mendelian genetic diseases across six priority domains: (I) Development, cognition, and mental health; (II) Feeding and growth; (III) Medication use and treatment history; (IV) Pain, sleep, and quality of life; (V) Adulthood; and (VI) Epilepsy. We also assessed in a similar manner follow-up reports appearing in the years following an initial report. These findings provided the impetus for, and guided the development of, the proposed new PHELIX (PHEnotype LIsting fiX) reporting guideline checklists.

## MATERIALS AND METHODS

### Systematic review

We utilized DistillerSR Version 2.35 for searching, screening, and data extraction (DistillerSR Inc, 2022; accessed January 2022 – January 2023). We identified all first reports of novel genetic conditions discovered through genotype-first ascertainment that result in multisystem and/or neurodevelopmental phenotypes, which were published in one of ten genetics journals (*American Journal of Human Genetics, American Journal of Medical Genetics Part A, Clinical Genetics, European Journal of Human Genetics, Genetics in Medicine, Genome Medicine, Human Molecular Genetics, Journal of Medical Genetics, Nature Genetics, PLoS Genetics*) during a 5-year period (1 January 2017 - 31 December 2021). The search executed on January 3, 2022, identified 3,243 articles (Supplementary Table 1; Supplementary Figure 1). Exclusion criteria were: (i) prenatal or neonatal lethal phenotype, (ii) case report or description of a single family, (iii) new gene for a known clinical syndrome (e.g., Joubert syndrome, Noonan syndrome), (iv) chromosome disorder with non-recurrent breakpoints that did not definitively implicate a specific gene, (v) potential genotype-phenotype expansion rather than a novel disorder. We also excluded large-scale gene discovery efforts in populations with common complex diseases and/or clinical testing laboratory cohort studies, where the *a priori* expectation for detailed phenotype descriptions was low. After both title and abstract screening and full-text review stages, n=200 reports describing 199 distinct monogenic conditions met the inclusion criteria (two reports of a novel condition were published at the same time; Supplementary Figure 1). For the subset of 25 genetic conditions first described in 2017, we performed an additional search using DistillerSR on June 1, 2022 (Supplementary Table 1) to identify subsequent published case descriptions (total n=95; Supplementary Figure 2). Reference review and additional internet searching did not identify any other “follow-up” reports.

**Table 1.**
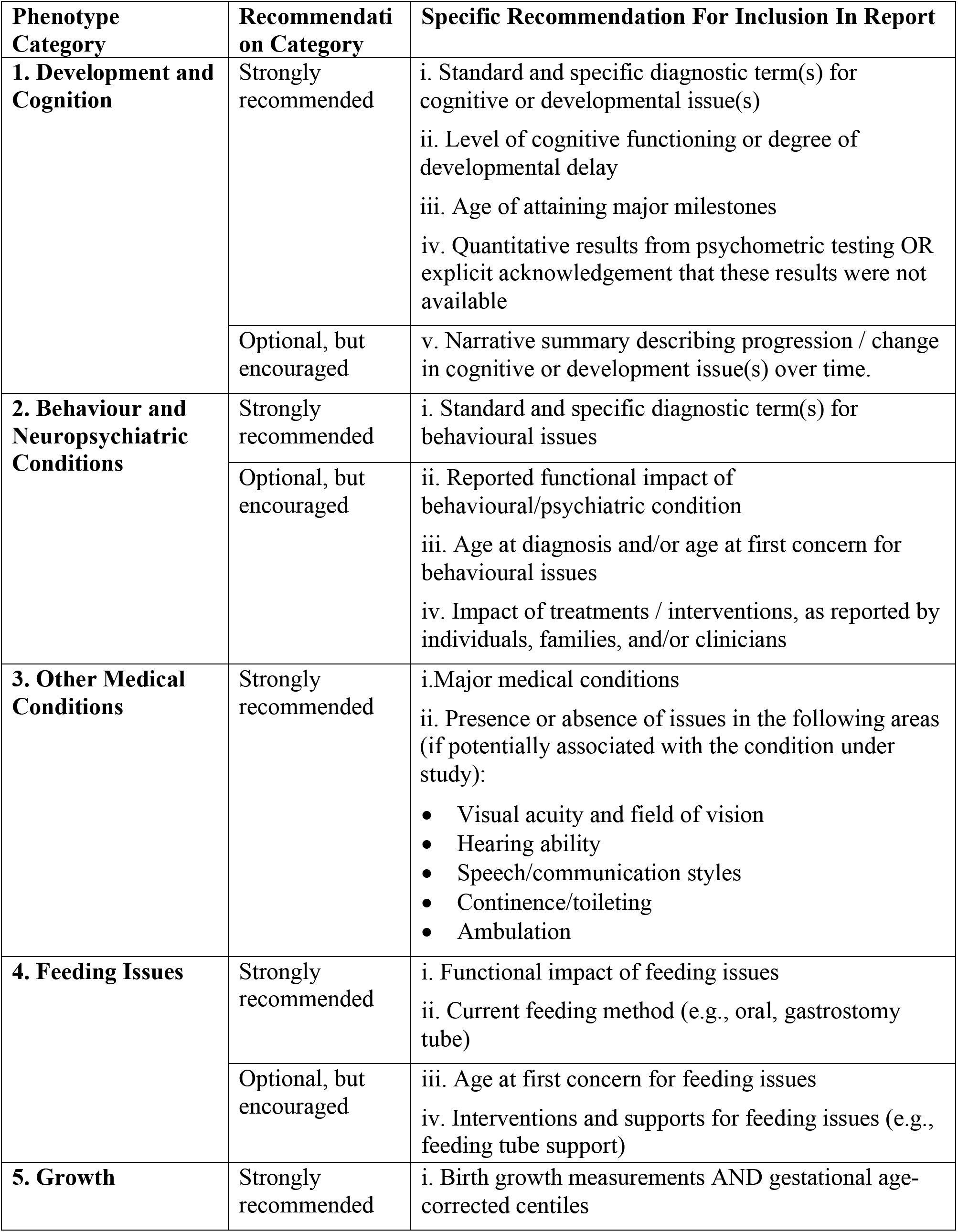
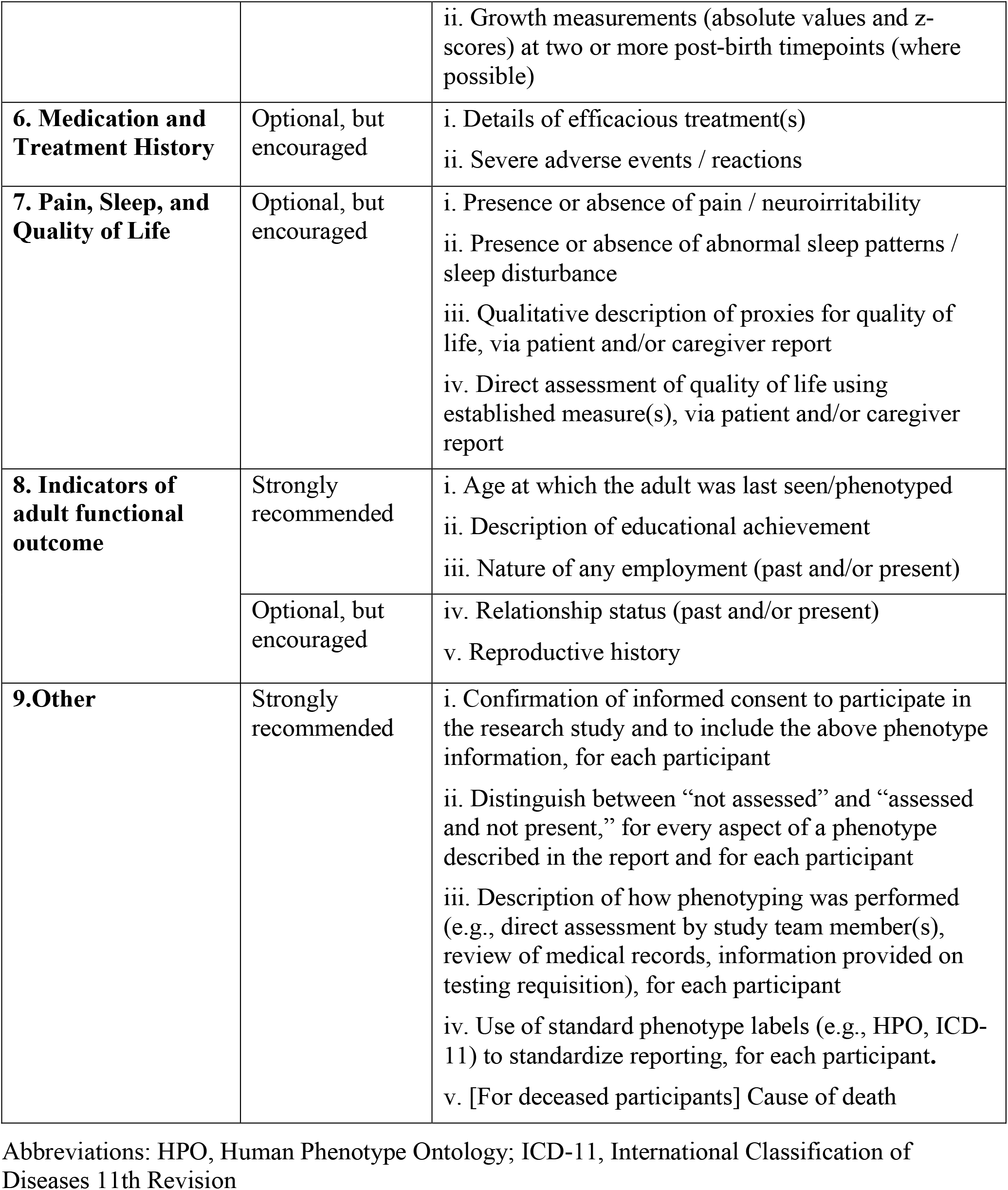
Phenotype reporting checklist (PHELIX_General version 1.0).

**Figure 1.**
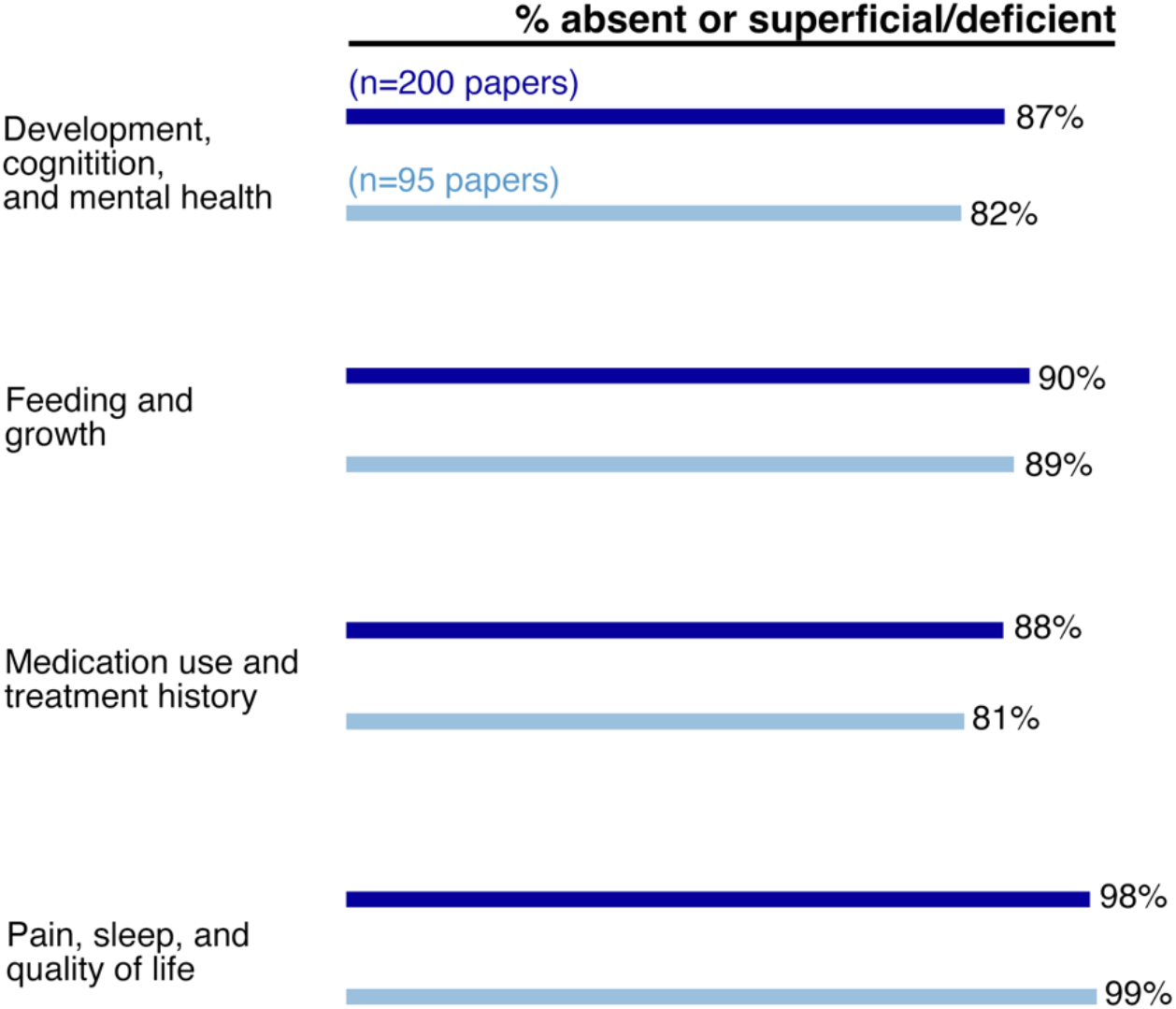
Global assessments of the quality of reporting of phenotype details germane to Domains I-IV. Dark blue = initial reports (n=200 papers), and light blue = follow-up reports regarding the 25 genetic conditions initially described in 2017 (n=95 papers). There were no significant differences in the distribution of overall quality ratings between the initial and follow-up reports, for any of the domains (Fisher’s exact tests, p>0.05). See text for details.

**Figure 2.**
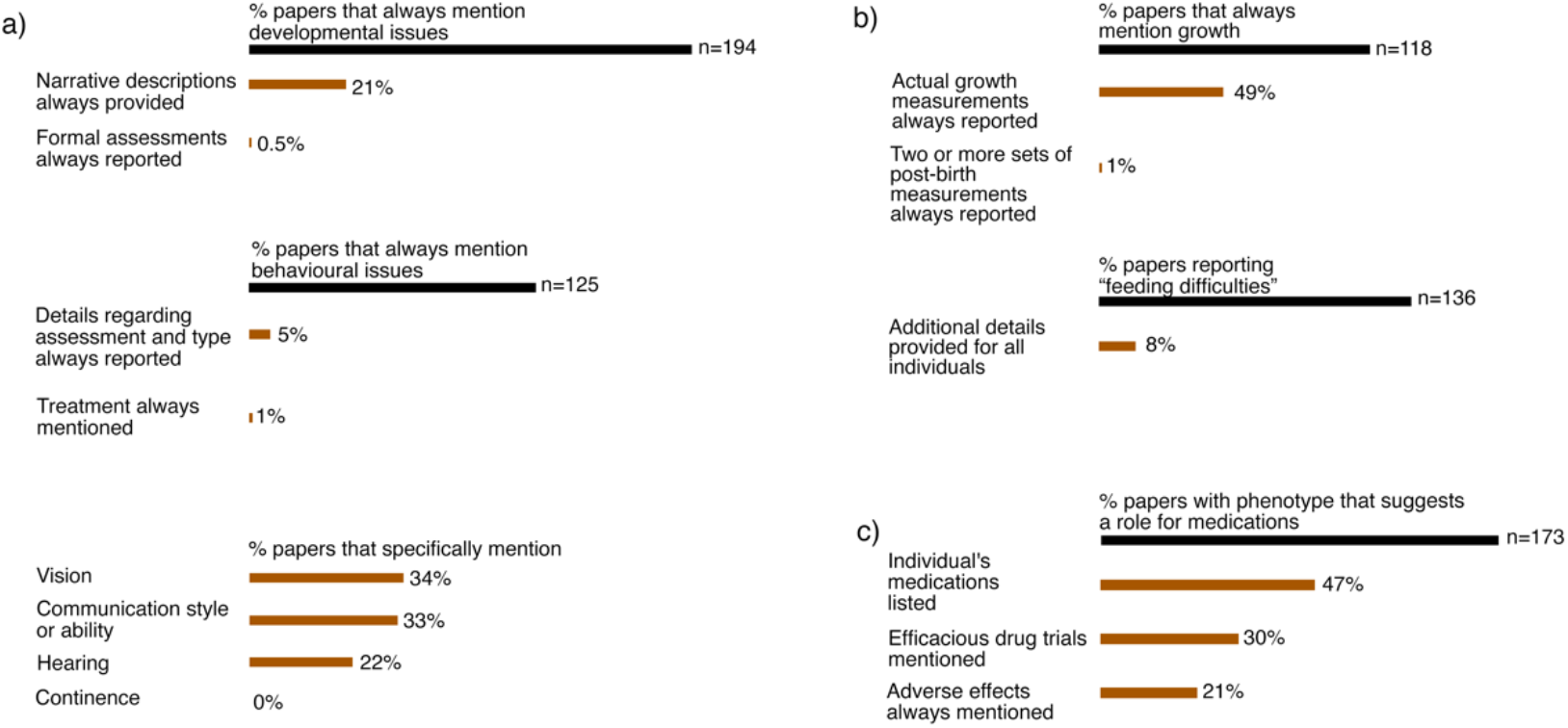
Selected item-level findings from a systematic review of phenotype reporting in n=200 descriptions of novel genetic syndromes. Domain I, (b) Domain II, (c), Domain III. See Supplementary Table 3 for details.

### Data extraction and analysis

For each published article (n=295), study team members (authors A.A., A.J., A.P., M.Y.F.) adjudicated the phenotype data pertaining to six priority domains [(I) Development, cognition, and mental health; (II) Feeding and growth; (III) Medication use and treatment history; (IV) Pain, sleep, and quality of life; (V) Adulthood; and (VI) Epilepsy] using a custom designed data extraction form (Supplementary Tables 3, 5, 6). Domains I-VI were included based on the study team’s clinical experience and review of the aforementioned published expert opinions, survey data, reviews, and data from patient and family focus groups.^14-24^ Data for Domains V-VI were extracted separately from Domains I-IV. The total 46-item form was developed by members of the study team (authors A.A., A.J., C.D., N.J., D.B., G.C.) with clinical expertise in medical genetics, psychiatry, development, general paediatrics, paediatric palliative care, paediatric complex care, and paediatric hospitalist medicine. Each domain was associated with multiple issue-specific items. A separate overall qualitative assessment of reporting quality (“strong”, “adequate”, “superficial/deficient”, “absent”, or “not applicable”) was also assigned for Domains I-IV. Descriptive statistics were calculated using Microsoft Excel.

### Development of phenotype reporting guidelines through a modified Delphi process

Medical experts from member institutions of the International Precision Child Health Partnership (IPCHiP) participated in a modified Delphi process.^25^ IPCHiP institutions included: Murdoch Children’s Research Institute/Royal Children’s Hospital (Melbourne, Australia), The Hospital for Sick Children (SickKids®; Toronto, Ontario, Canada), University College London/Greater Ormond Street Hospital (London, United Kingdom), and Boston Children’s Hospital (Massachusetts, USA).^26,27^ At the suggestion of the original study team members, additional expertise was sought in: (i) neuropsychological assessment and cognitive phenotyping (via Seaver Autism Center for Research and Treatment; New York, USA),^28-30^ and (ii) adult phenotyping (via University Health Network; Toronto, Ontario, Canada).^31,32^ Authors J.C., L.D., P.G., T.L., P.S., Z.S., J.A.S.V., C.D., N.J., D.B. contributed to the initial refinement of guidelines for Domains I-V. We sent out three electronic surveys to the above authors (minimum engagement rate >50%) over a five-month period to define and prioritize the reporting criteria. We then hosted an online meeting that incorporated independent voting on inclusion/exclusion of each draft item. The meeting was recorded, to allow for asynchronous viewing by those expert volunteers who were unable to attend in real-time. Authors V.C., A.D., K.H., N.S.Y.L., A.T., A.P., K.W. contributed to the initial refinement of guidelines for Domain VI (Epilepsy). Similarly, we used a series of two electronic surveys to define and prioritize the reporting criteria. All authors reviewed, revised, and ultimately approved the reporting guideline checklists for Domains I-VI reported herein. The guideline checklists will be uploaded to the EQUATOR (Enhancing the QUAlity and Transparency Of health Research) Network website (https://www.equator-network.org/) as the PHELIX_General (Table 1) and the PHELIX_Epilepsy checklists (Table 2).

**Table 2.**
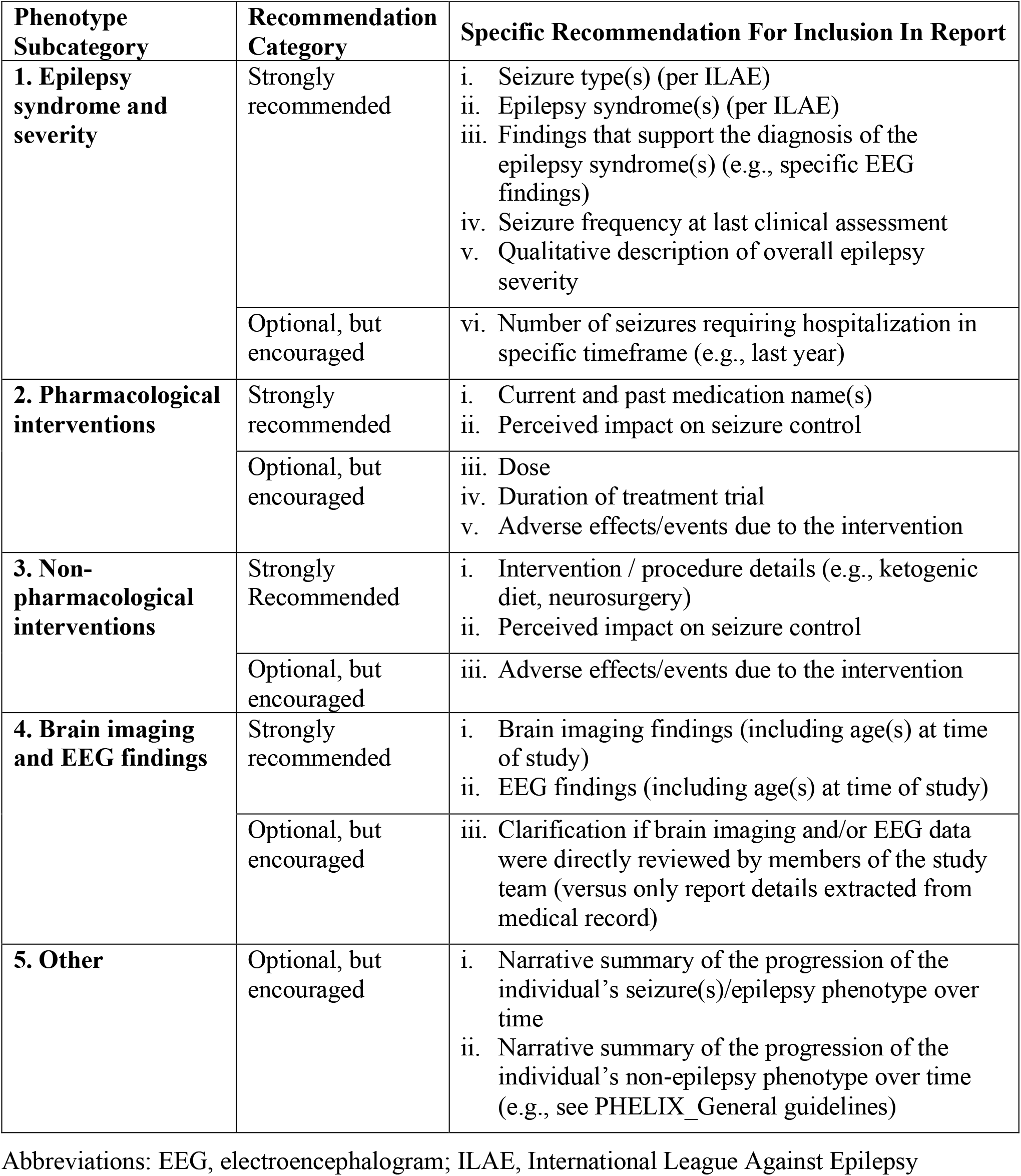
Phenotype reporting recommendations for unprovoked seizures/epilepsy in individuals with newly described multisystem and/or neurodevelopmental Mendelian disorders (PHELIX_Epilepsy version 1.0).

## RESULTS

### Contemporary descriptions of new syndromes are often lacking in phenotype details

The 200 reports of 199 newly discovered genetic disorders included phenotype descriptions for a total of 1,856 study participants (median: 7/report, range 2-42). Features of the reports (year and journal of publication) and of the participants (age) are summarized in Supplementary Tables 2-3). The overall qualitative assessment of reporting was deemed “superficial/deficient” or “absent” in 87% (Domain I: Development, cognition, and mental health) to 98% (Domain IV: Pain, sleep, and quality of life) of papers (Figure 1). Five (2.5% of 200) reports were deemed “strong” in any single domain (pertaining to the genetic conditions associated with variants in the genes *ADARB1*^33^, *GNAI1*^34^, *NCAPG2*^35^, *PCDHGC4*^36^, and *SPTBN1*^37^). No reports were deemed “strong” in their reporting across each of Domains I-IV. The year and journal of publication were not associated with overall quality assessment of phenotype reporting (data not shown).

Item-specific data supported the overall qualitative assessments of reporting quality (Figure 2, Supplementary Table 3, Supplementary Figures 3-4). While 97% of papers mentioned developmental concerns in study participants, 21% provided details about cognitive abilities for all the participants and a sole paper^38^ reported results from formal cognitive assessments for all participants (Figure 2a, Supplementary Table 4). A common issue was that individuals were identified as having “developmental delay” without further elaboration. Similarly, of the papers that reported neuropsychiatric and behavioural issues in study participants, less than 5% of papers provided details for all participants regarding type/diagnosis, symptom severity, and/or nature of the assessments (Figure 2a, Supplementary Table 3). Of the papers that reported on the presence of feeding difficulties (Figure 2b), 8% consistently reported on the type/nature of feeding issues and current means of feeding (Figure 2b, Supplementary Table 3). Growth parameters at birth were often reported, but 6% of papers reported on two or more growth measurements post-birth to allow for assessment of growth trajectories (Figure 2b, Supplementary Table 4). Nearly half of all papers made no mention of participants’ medications or treatment trials, or of the absence thereof (Figure 2c). The presence or absence of adverse effects of treatments were explicitly mentioned in just 21% of reports (Figure 2c).

Domain V (Adulthood) was assessed in the subset of reports that included at least one adult individual (n=63; adult defined as age >=18 years) (Supplementary Figure 1; Supplementary Table 6). Domain VI (Epilepsy) was assessed in the subset of reports that included at least one study participant with seizures/epilepsy (n=85) (Supplementary Figure 1; Supplementary Table 7). Consistent with the findings regarding Domains I-IV, most items were inconsistently or never reported (Supplementary Figures 3-4). For example, papers rarely described proxies for adult functioning such as educational achievement or employment, nor the anti-seizure treatments for individuals with epilepsy.

### Follow-up reports do not consistently address initial gaps in phenotype descriptions

Regarding the 25 genetic conditions first described in 2017, the 95 “follow-up” reports included phenotype descriptions for an additional 334 study participants (median: 1 per report, range 1-25). The overall qualitative assessment of reporting was similarly classified as “absent” or “superficial/deficient” in 81% (Domain III: Medication use and treatment history) to 99% (Domain IV: Pain, sleep, and quality of life) of papers (Figure 1, Supplementary Table 4), with no significant differences between the original and the follow-up reports. Eleven reports were deemed “strong” in any single domain (pertaining to the genetic conditions associated with variants in the eight genes *CAMK2A*^39^, *CAMK2B*^39,40^, *DHX30*^41^, *OTUD6B*^42^, *PPP3CA*^43^, *UBTF*^44,45^, *WDR26*^46,47^, and *YY1*^48^). No reports were deemed “strong” in their reporting across each of domains I-IV. Item-specific data are summarized in Supplementary Table 4.

### Consensus phenotype reporting guidelines

Guideline checklists to enhance the reporting of phenotype data for ultra-rare genetic conditions were developed through a modified Delphi process^25^ and informed by the findings above (Supplementary Table 8). Specifically, items were included based on their superficial/deficient reporting in the literature to date, and on the recommendations of expert collaborators as being data that are both important to capture and feasible to obtain by researchers. The finalized checklist of 33 items across 9 categories is presented in Table 1 (PHELIX_General). To showcase how these guidelines could be expanded over time, additional items specific to epilepsy phenotype reporting are listed in Table 2 (PHELIX_Epilepsy). Extended versions with examples are provided in Supplementary Tables 9-10.

## DISCUSSION

Our results reveal that phenotype information relevant to clinical management, genetic counseling, and the stated priorities of patients and families, is lacking for many newly described genetic diseases. Although most published reports acknowledged the key phenotype domains assessed, few original or follow-up reports included clinically relevant details. To address this issue, we propose reporting guideline checklists for use by researchers and journals. Use of these guidelines could improve phenotype reporting in the era of genotype-first and matchmaker service-driven reports of novel syndromes. Decision making about precision genetic or other therapy development, including the potential for n=1 trials, may be contingent on our understanding (or lack thereof) of the natural history of a given ultra-rare genetic disease.^10,11,49^

Reasons for under-reporting phenotype data are likely multiple and complex. First, these data may not be readily available to the referring clinician or laboratory collaborators, and “phenotyping is hard,”^50^ especially for older individuals with extensive past histories. Review of lifetime medical records, and/or a brief, targeted interview with patients and/or their caregiver(s), should be sufficient to gather most of the information outlined in Tables 1-2. Second, these data may not be requested by the coordinating research team that is leading the publication effort. In our experience, many groups design their own data collection forms that ask for no or only general details regarding issues outside of that group’s specific phenotype(s) of interest. Third, unlike for example DNA sequencing methods, there are no defined minimum reporting standards for phenotyping to guide peer reviewers and journal editors. Finally, there may be a belief that phenotype reporting in initial descriptions of novel genetic diseases is less important than establishing an association between variation in the gene and (any) disease phenotype. Although the hope may be that future reports will then describe many more individuals and include detailed phenotype data, we did not find evidence that this is consistently happening in practice in a timely manner.

We recognize several limitations of our review and guideline development methods. We selected only ten top-tier genetics journals for our systematic review. The generalizability of our findings to reports published in other specialty-specific or organ system-specific journals is unclear. Out of necessity given the lack of validated tools, we created a new data collection questionnaire to assess the reporting of phenotype data and relied on subjective assessments from raters for some items. We selected broad phenotype domains based on our combined clinical experiences and the published literature. Ours was a paediatrics-focused effort, reflecting the phenotypes that are currently driving most Mendelian gene discovery efforts. Other groups may develop and add-on reporting criteria for additional specific phenotype elements, as we did for epilepsy, and continue to refine the general adult phenotype elements (Table 2). We also restricted our initial focus to cross-sectional reporting, and additional guidance will be needed for evaluating within-individual natural history. Finally, our reporting guidelines have not yet been applied prospectively to assess feasibility and utility.

We propose minimum standards for phenotype descriptions of putative novel disorders with multisystem manifestations and/or a neurodevelopmental component in children. Further refinement of our proposed reporting guidelines is an important consideration. Collecting additional input from key stakeholders (e.g., rare disease organizations, journal editors) should be coupled with attempts to better integrate technologies for systematic phenotype collection and data sharing.^21,51^ Improved reporting of phenotype aspects like craniofacial morphology (“dysmorphic features”)^52^ could help with interpreting variants of uncertain significance and assessing phenotypic “fit”.^53,54^ The aim of the PHELIX guideline checklists is to decrease the variability in the consistency of phenotyping and description of findings, and thereby enhance the ongoing clinical care of individuals with genetic conditions.

## ACKNOWLEDGMENTS

Funding was provided by the SickKids Research Institute, the Canadian Institutes of Health Research (Funding Reference Number: PJT186240), and the University of Toronto McLaughlin Centre. IPCHiP is supported in part by SickKids Foundation donors including Jamie and Patsy Anderson and the Feiga Bresver Academic Foundation. D.B. also acknowledges the support of her research time at Holland Bloorview by the Arthur Family Foundation. J.A.S.V. holds the SickKids Psychiatry Associates Chair in Developmental Psychopathology at The Hospital for Sick Children. Research conducted at the Murdoch Children’s Research Institute was supported by the Victorian Government’s Operational Infrastructure Support Program. The Chair in Genomic Medicine awarded to J.C. is generously supported by The Royal Children’s Hospital Foundation. These funders played no role in study design, data collection, analysis and interpretation of data, or the writing of this manuscript.

## CRediT AUTHOR STATEMENT

Conceptualization: D.B., G.C.

Methodology: A.A., A.J., C.D., N.J., D.B., G.C.

Software: N/A

Validation: A.A., A.J., A.P., M.Y.F., G.C.

Formal analysis: A.A.

Investigation: A.A., A.J., A.P., M.Y.F., V.C., A.D., K.H., N.S.Y.L., A.T., A.P., K.W., A.S.B., J.C.,

L.D., P.G., T.L., P.S., Z.S., J.A.S.V., C.D., N.J., D.B., G.C.

Resources: G.C.

Data curation: N/A

Writing – original draft: A.A., A.J., D.B., G.C.

Writing – review & editing: A.P., M.Y.F., V.C., A.D., K.H., N.S.Y.L., A.T., A.P., K.W., A.S.B., J.C., L.D., P.G., T.L., P.S., Z.S., J.A.S.V., C.D., N.J.

Visualization: A.A., A.J. Supervision: L.D., P.G., G.C.

Project administration: A.A., A.J., D.B., G.C. Funding acquisition: G.C.

## DATA AVAILABILITY STATEMENT

The datasets analysed during the current study are available from the corresponding authors on reasonable request.

