## Supplemental Figures 1-4 and Supplemental Tables 1-10 for "Gaps in the phenotype descriptions of ultra-rare genetic conditions: review and multicenter consensus reporting guidelines"

**SUPPLEMENTARY FILE FOR:** Gaps in the phenotype descriptions of ultra-rare genetic conditions: review and multi-center consensus reporting guidelines

**TABLE OF CONTENTS**

Supplementary Figure 1…………………………………………………………...………………2
Supplementary Figure 2…………………………………………………………...……………3
Supplementary Figure 3…………………………………………………………...…………4
Supplementary Figure 4…………………………………………………………...………5
Supplementary Table 1……………………………………………………………...……6
Supplementary Table 2…………………………………………………………………....…7
Supplementary Table 3……………………………………………………………………...8
Supplementary Table 4……………………………………………………………………9
Supplementary Table 5…………………………………………………………………..14
Supplementary Table 6………………………………………………………………….15
Supplementary Table 7…………………………………………………………………..17
Supplementary Table 8…………………………………………………………………..19
Supplementary Table 9…………………………………………………………………..20
Supplementary Table 10…………………………………………………………………23

**Supplementary Figure 1. PRISMA flow diagram for primary search.**
See main text for details.

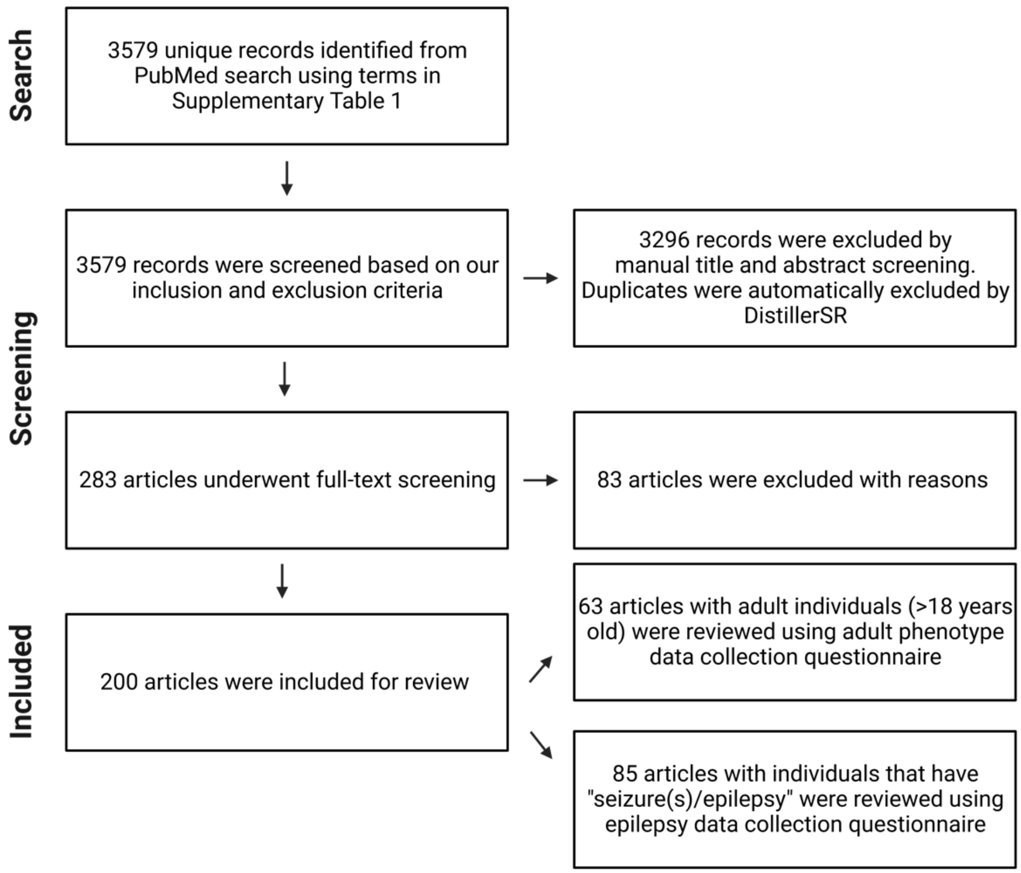

**Supplementary Figure 2. PRISMA flow diagram for secondary search.**
See main text for details.

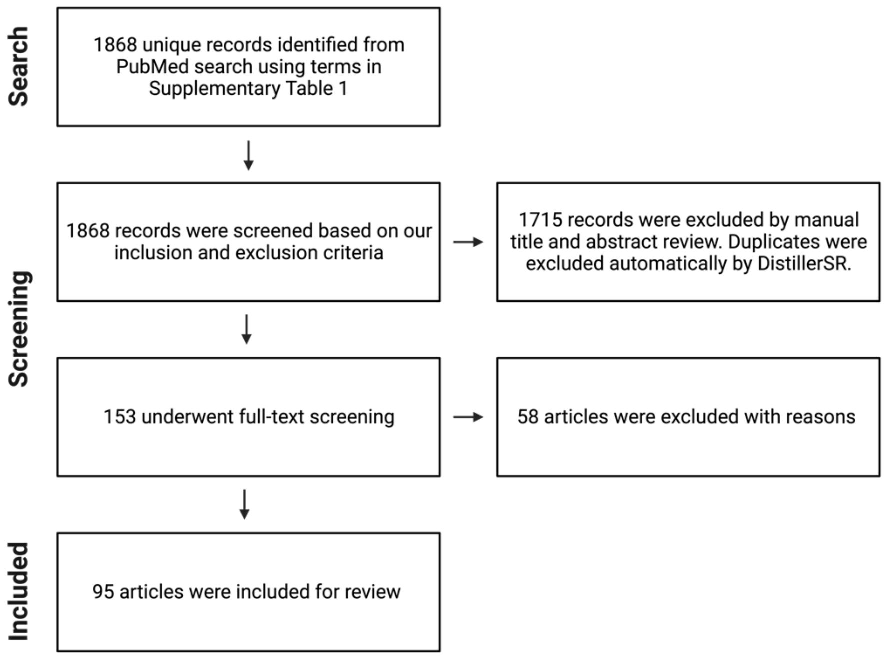

**Supplementary Figure 3. Lollipop chart summarizing reporting of adulthood-related aspects of phenotype in n=63 reports of novel genetic disorders.**
See main text for details.

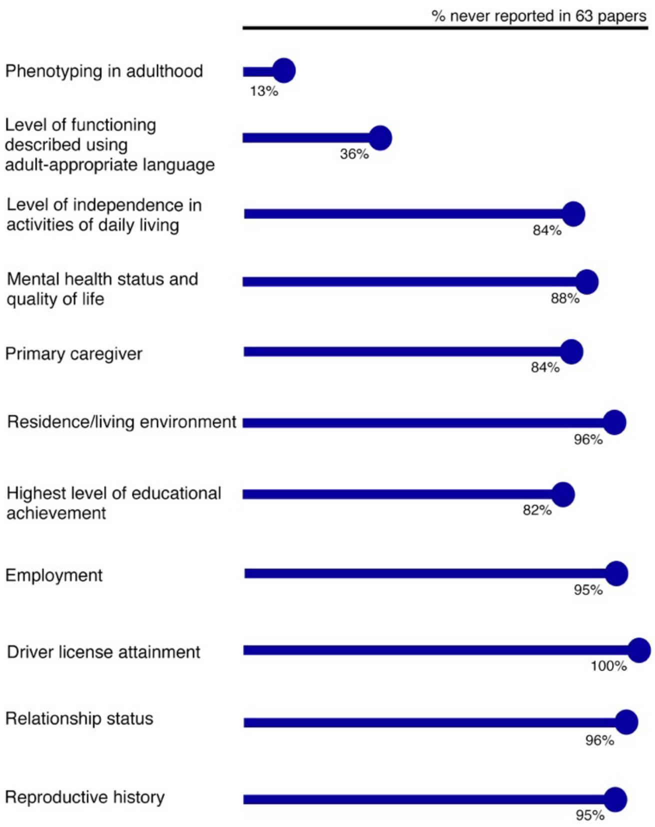

**Supplementary Figure 4. Lollipop chart summarizing reporting of seizure/epilepsy-related aspects of phenotype in n=85 reports of novel genetic disorders.**
See main text for details.

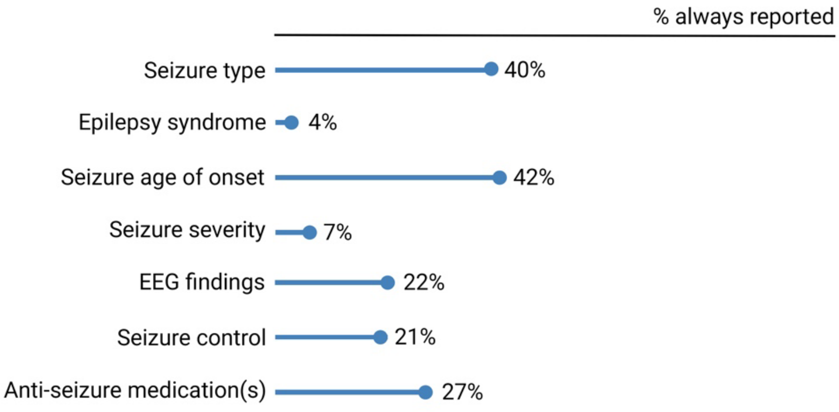

**Supplementary Table 1. DistillerSR search details.**

| Search Type | Search Date | Search Terms |
|---|---|---|
| Initial reports | January 3, 2022 | (Nature Genetics[journal] OR American Journal of Human Genetics[journal] OR Genome Medicine[journal] OR PloS Genetics[journal] OR Genetics in Medicine[journal] OR Human Molecular Genetics[journal] OR Journal of Medical Genetics[journal] OR American Journal of Medical Genetics Part A[journal] OR European Journal of Human Genetics[journal] OR European Journal of Medical Genetics[journal] OR Journal of Human Genetics[journal] OR Clinical Genetics[journal]) AND 2017:2021[dp] AND (new[title] OR novel[title] OR associated[title] OR cause[title] OR causes[title]) |
| Follow-up reports | June 1, 2022 | OTUD6B OR PPM1D OR ISCA1 OR PSMD12 OR PLAA OR ZNF462 OR YY1 OR NKX6-2 OR KCNQ5 OR WDR26 OR LIPT2 OR UBTF OR SCAPER OR CDK10 OR HIST1H4C OR C1QBP OR BPTF OR PPP3CA OR FDXR OR DHX30 OR CAMK2A OR CAMK2B OR H4C3 OR GPAA1 OR RAB11B |

**Supplementary Table 2. Publication details for the n=200 reports of novel genetic disorders included in this study.**
See main text and Supplementary Figure 1 for details. There were no significant differences in the distribution of qualitative assessments of phenotype Domains by year or by journal (data not shown).

| **Year of Publication** | **Number of Reports** |
|---|---|
| 2017 | 24 |
| 2018 | 38 |
| 2019 | 57 |
| 2020 | 37 |
| 2021 | 44 |
| **Journal** | |
| *American Journal of Human Genetics* | 134 |
| *Clinical Genetics* | 4 |
| *European Journal of Human Genetics* | 9 |
| *Genetics in Medicine* | 19 |
| *Genome Medicine* | 1 |
| *Human Molecular Genetics* | 15 |
| *Journal of Medical Genetics* | 11 |
| *Nature Genetics* | 5 |
| *PLoS Genetics* | 2 |

**Supplementary Table 3. Ages of research participants in 200 reports of novel genetic disorders included in this study.**

| Age range in years | Number of individuals (% of n=1856) |
|---|---|
| <2 | 203 (10.9%) |
| 2-7 | 651 (35.1%) |
| 8-17 | 622 (33.5%) |
| >17 | 263 (14.2%) |
| Not reported | 117 (6.3%) |

**Supplementary Table 4. Phenotype extraction form data for Domains I-IV for the initial and follow-up reports.**

| | Initial Reports (n=200) | Follow-up Reports (n=95) |
|---|---|---|
| **DOMAIN I: DEVELOPMENT, COGNITION, AND MENTAL HEALTH** | | |
| 1. Are development and/or cognition issues mentioned as a feature? (e.g., global developmental delay / intellectual disability / encephalopathy/ cognitive impairment / motor impairment / social communication deficits) | Yes: 96.5%<br>No: 3.5% | Yes: 87.4%<br>No: 12.6% |
| a. Are qualifiers used? (e.g., mild, moderate, severe, profound) | Always: 42%<br>Sometimes: 40.4%<br>Never:17.6% | Always: 43.4%<br>Sometimes: 19.3%<br>Never: 37.3% |
| b. Are narrative descriptions of development and cognition provided? | Always: 21.2%<br>Sometimes: 53.9%<br>Never: 24.9% | Always: 61.4%<br>Sometimes: 12.1%<br>Never: 26.5% |
| c. Were formal assessments performed? (e.g., WISC, WAIS, Bayley) | Always: 0.5%<br>Sometimes: 37.5%<br>Never: 62% | Always: 19.3%<br>Sometimes: 16.8%<br>Never: 63.9% |
| 2. Are behavioural and/or psychiatric issues mentioned as a feature? | Yes: 62.5%<br>No: 37.5% | Yes: 50%<br>No: 50% |
| a. Are details given regarding assessment, type, and severity? | Always: 4.8%<br>Sometimes: 75.2%<br>Never: 2% | Always: 13%<br>Sometimes: 13%<br>Never: 74% |
| b. Is age of onset specified? | Always: 6.4%<br>Sometimes: 24.8%<br>Never: 68.8% | Always: 6.5%<br>Sometimes: 2.2%<br>Never: 91.3% |
| c. Is treatment mentioned? | Always: 0.8%<br>Sometimes: 12.8%<br>Never: 86.4% | Always: 6.5%<br>Sometimes: 2.2%<br>Never: 91.3% |
| 3. Is adaptive functioning or a relevant proxy/component described? (i.e., continence, hearing, vision, | Yes: 95%<br>No: 5% | Yes: 76.8%<br>No: 23.2% |

| | | |
|---|---|---|
| technology dependence, communication) | | |
| a. Is continence mentioned? | Always: 0%<br>Sometimes: 11.8%<br>Never: 88.2% | Always: 4.1%<br>Sometimes: 5.4%<br>Never: 90.5% |
| b. Is vision mentioned? | Always: 34.2%<br>Sometimes: 40.5%<br>Never: 25.3% | Always: 56.8%<br>Sometimes: 17.6%<br>Never: 25.7% |
| c. Is hearing mentioned? | Always: 22.1%<br>Sometimes: 38.4%<br>Never: 39.5% | Always: 21.6%<br>Sometimes: 16.2%<br>Never: 62.2% |
| d. Is communication style / ability mentioned? | Always: 33.2%<br>Sometimes: 57.9%<br>Never: 8.9% | Always: 48.6%<br>Sometimes: 21.6%<br>Never: 29.8% |
| 4. Is adaptive functioning described in quantitative terms? (e.g., ABAS or Vineland)? | Always: 0%<br>Sometimes: 7.4%<br>Never: 92.6% | Always: 10.8%<br>Sometimes: 4.1%<br>Never: 85.1% |
| 5. Is technology (assistive devices) use mentioned? | Yes: 56.6%<br>No: 43.3% | Yes: 33.3%<br>No: 66.7% |
| a. Is need for breathing tube mentioned? | Always: 0%<br>Sometimes: 17%<br>Never: 83% | Always: 3.8%<br>Sometimes: 15.4%<br>Never: 80.8% |
| b. Is need for wheelchair mentioned? | Always: 0%<br>Sometimes: 32%<br>Never: 68% | Always: 0%<br>Sometimes: 24%<br>Never: 76% |
| c. Is need for feeding tube mentioned? | Always: 8.4%<br>Sometimes: 74.8%<br>Never: 16.8% | Always: 24%<br>Sometimes: 48%<br>Never: 28% |
| 6. Overall rating for [this domain] | Absent: 1.5%<br>Superficial/Deficient: 85.5%<br><br>Adequate: 12.0%<br>Strong: 1% | Absent: 5.3%<br>Superficial/Deficient: 76.8%<br><br>Adequate: 12.6%<br>Strong: 5.2% |
| **DOMAIN II: FEEDING AND GROWTH** | | |

| | | |
|---|---|---|
| 7. Are "feeding difficulties" mentioned as a feature? | Always: 18%<br>Sometimes: 50%<br>Never: 32% | Always: 29.5%<br>Sometimes: 20%<br>Never: 50.5% |
| a. Are any additional qualifiers or contextual details provided at all (e.g., age at onset, severity, and/or necessary supports)? | Always: 8.2%<br>Sometimes: 74.6%<br>Never: 17.2% | Always: 18.8%<br>Sometimes: 12.5%<br>Never: 68.7% |
| 8. Is growth mentioned? | Always: 59%<br>Sometimes: 33.5%<br>Never: 7.5% | Always: 69.5%<br>Sometimes: 2.1%<br>Never: 28.4% |
| a. Are the actual growth measurements and corresponding ages provided? (not just percentile ranges) | Always: 49.2%<br>Sometimes: 37%<br>Never: 13.5% | Always: 62.3%<br>Sometimes: 17.4%<br>Never: 20.3% |
| b. Are measurements provided for two or more time points (outside of birth parameters)? | Always: 1.1%<br>Sometimes: 4.9%<br>Never: 94% | Always: 7.3%<br>Sometimes: 5.8%<br>Never: 86.9% |
| 9. Overall rating for [this domain] | Absent: 3%<br>Superficial/ Deficient: 87%<br>Adequate: 9%<br>Strong: 1 % | Absent: 24.2%<br>Superficial/ Deficient: 64.2%<br>Adequate: 10.5%<br>Strong: 1.1% |
| **DOMAIN III: MEDICATION USE AND TREATMENT HISTORY** | | |
| 10. Are there any aspects of the phenotype that suggest a possible role for medication(s)? (e.g., seizures, "behavioural problems", aggression, sleep disturbance, etc) | Yes: 86.5%<br>No: 13.5% | Yes: 66.3%<br>No: 33.7% |
| 11. Are any of an individual's prescription medications listed? | Always: 8.5%<br>Sometimes: 38%<br>Never: 53.5% | Always: 20%<br>Sometimes: 10.5%<br>Never: 69.5% |
| 12. Is the total number of prescribed medications per participant listed? | Always: 5.5%<br>Sometimes: 15.5%<br>Never: 79% | Always: 2.1%<br>Sometimes: 1.1%<br>Never: 96.8% |
| 13. Are adverse drug events (or lack thereof) mentioned? | Always: 0%<br>Sometimes: 5%<br>Never: 95% | Always: 6.3%<br>Sometimes: 2.1%<br>Never: 91.6% |
| 14. Are successful drug trials mentioned? (i.e., positive responses to medications) | Always: 4%<br>Sometimes: 25.5%<br>Never: 70.5% | Always: 15.8%<br>Sometimes: 9.5%<br>Never: 74.7% |

| | | | |
|---|---|---|---|
| 15. | Are unsuccessful drug trials mentioned? (i.e., negative responses to medications) | Always: 2.5%<br>Sometimes: 18.5%<br>Never: 79% | Always: 15.8%<br>Sometimes: 8.4%<br>Never: 75.8% |
| 16. | Were other treatments discussed? (e.g., dietary, surgical; excluding nutritional interventions indicated previously) | Yes: 48.5%<br>No: 51.5% | Yes: 37.9%<br>No: 62.1% |
| 17. | Overall rating for [this domain] | Absent: 35.5%<br>Superficial /Deficient: 52.5%<br>Adequate: 11%<br>Strong: 1% | Absent: 48.4%<br>Superficial /Deficient: 32.6%<br>Adequate: 12.4%<br>Strong: 6.3% |
| **DOMAIN IV: PAIN, SLEEP, AND QUALITY OF LIFE** | | | |
| 18. | Is the presence/absence of pain and neuro-irritability mentioned? | Always: 0%<br>Sometimes: 18%<br>Never: 82% | Always: 6.3%<br>Sometimes: 7.4%<br>Never: 86.3% |
| 19. | Is quality of life characterized using an established scale/questionnaire/assessment? | Always: 0%<br>Sometimes: 2.5%<br>Never: 97.5% | Always: 0%<br>Sometimes: 0%<br>Never: 100% |
| 20. | Is the presence/absence of sleep problems mentioned? | Always: 4%<br>Sometimes: 35%<br>Never: 61% | Always: 7.4%<br>Sometimes: 11.6%<br>Never: 81% |
| 21. | Is mortality mentioned? | Yes: 27.5%<br>No: 72.5% | Yes: 14.7%<br>No: 85.3% |
| | a. Are age at death and cause of death described? | Always: 51.9%<br>Sometimes: 38.9%<br>Never: 9.2% | Always: 80%<br>Sometimes: 13.3%<br>Never: 6.7% |
| | b. Is palliative care involvement mentioned? | Yes: 1.9%<br>No: 98.1% | Yes: 0%<br>No: 100% |
| 22. | Is the presence/absence of sialorrhea mentioned? | Always: 0%<br>Sometimes: 8.5%<br>Never: 91.5% | Always: 6.3%<br>Sometimes: 7.4%<br>Never: 86.3% |
| 23. | Is the presence/absence of recurrent aspirations mentioned? | Always: 1%<br>Sometimes:12%<br>Never: 87% | Always: 2.1%<br>Sometimes:4.2%<br>Never: 93.7% |

| | | | |
|---|---|---|---|
| 24. | Are the presence/absence of tone abnormalities mentioned? (e.g., hypotonia, hypertonia, dystonia) | Always: 59.5%<br>Sometimes: 28.5%<br>Never: 12% | Always: 47.4%<br>Sometimes: 15.8%<br>Never: 36.8% |
| 25. | Overall rating for [this domain] | Absent: 6%<br>Superficial/Deficient: 92%<br>Adequate: 2%<br>Strong: 0% | Absent: 26.3%<br>Superficial/Deficient: 72.6%<br>Adequate: 1.1%<br>Strong: 0% |

**Supplementary Table 5. PMIDs of the n=295 papers reviewed in this study.**

| | | | | | | | |
|---|---|---|---|---|---|---|---|
| 28007986 | 29276004 | 30335141 | 30905399 | 31495489 | 32413283 | 33442026 | 34145223 |
| 28130356 | 29290337 | 30343942 | 30929739 | 31509304 | 32430360 | 33473207 | 34180050 |
| 28135719 | 29300972 | 30343943 | 30941876 | 31564433 | 32442410 | 33506510 | 34211179 |
| 28343629 | 29304374 | 30359776 | 30976112 | 31585109 | 32527956 | 33513338 | 34230638 |
| 28343630 | 29304375 | 30364145 | 30976113 | 31587868 | 32543299 | 33522091 | 34244665 |
| 28356563 | 29388673 | 30388400 | 30982608 | 31607425 | 32593294 | 33531666 | 34346499 |
| 28388435 | 29394991 | 30388402 | 31006512 | 31630790 | 32600977 | 33632302 | 34354232 |
| 28413018 | 29427787 | 30401460 | 31031012 | 31735293 | 32627382 | 33638881 | 34355505 |
| 28513610 | 29432562 | 30401461 | 31034465 | 31785787 | 32652806 | 33675273 | 34379057 |
| 28575647 | 29478781 | 30421579 | 31036918 | 31838722 | 32694869 | 33711248 | 34385670 |
| 28575651 | 29483653 | 30455226 | 31069901 | 31883644 | 32707086 | 33742450 | 34411328 |
| 28669405 | 29560374 | 30481285 | 31079897 | 31916397 | 32721402 | 33743206 | 34413497 |
| 28686853 | 29576218 | 30487643 | 31079898 | 31924697 | 32730804 | 33782605 | 34419324 |
| 28692176 | 29656858 | 30500825 | 31079899 | 31929335 | 32738225 | 33796307 | 34585832 |
| 28757203 | 29656859 | 30503518 | 31089205 | 31931739 | 32779182 | 33811806 | 34587489 |
| 28777933 | 29656860 | 30517966 | 31092906 | 31949313 | 32820033 | 33824500 | 34618373 |
| 28794130 | 29726930 | 30526862 | 31124279 | 32004446 | 32822602 | 33847457 | 34703884 |
| 28837161 | 29758292 | 30526868 | 31147255 | 32004679 | 32875707 | 33864376 | 34707297 |
| 28866611 | 29767723 | 30549423 | 31155615 | 32030560 | 32885237 | 33909591 | 34715011 |
| 28886341 | 29808498 | 30561111 | 31178128 | 32031333 | 32891193 | 33909990 | 34729769 |
| 28920961 | 29907796 | 30580808 | 31192531 | 32092383 | 32924626 | 33909992 | 34804873 |
| 28934986 | 29961568 | 30595371 | 31204009 | 32109420 | 33026538 | 33938912 | 34906456 |
| 28940097 | 29961569 | 30595372 | 31230721 | 32152250 | 33073849 | 33963760 | 34906466 |
| 28942965 | 29979980 | 30609410 | 31256876 | 32181568 | 33110267 | 33975400 | 34906501 |
| 28942966 | 30057030 | 30612693 | 31278393 | 32197073 | 33159882 | 33979636 | 35016835 |
| 28942967 | 30057031 | 30633344 | 31303265 | 32197074 | 33186545 | 33991472 | 35027293 |
| 28965846 | 30105122 | 30639322 | 31322726 | 32197075 | 33217309 | 34003581 | 35080150 |
| 28969374 | 30122539 | 30661771 | 31327508 | 32214227 | 33232675 | 34003604 | 35172867 |
| 29040572 | 30214071 | 30661772 | 31327510 | 32220290 | 33245860 | 34010604 | 35198003 |
| 29100085 | 30224647 | 30679813 | 31353023 | 32220291 | 33276377 | 34020708 | 35202563 |
| 29100089 | 30250212 | 30723319 | 31353024 | 32246862 | 33308444 | 34022130 | 35377796 |
| 29100095 | 30254215 | 30755392 | 31361404 | 32275884 | 33344382 | 34050707 | 35430327 |
| 29106825 | 30269814 | 30795918 | 31363758 | 32307552 | 33348459 | 34055682 | 35503477 |
| 29130579 | 30290153 | 30824121 | 31402090 | 32330417 | 33355653 | 34077761 | 35583973 |
| 29158550 | 30290154 | 30827496 | 31422817 | 32338762 | 33369188 | 34102099 | 35620293 |
| 29251763 | 30293988 | 30827498 | 31422819 | 32341456 | 33417887 | 34114225 | 35627197 |
| 29267967 | 30323019 | 30905398 | 31439720 | 32356556 | 33420346 | 34143952 | |

**Supplementary Table 6. Phenotype extraction form data for Domain V for the initial reports describing adults.**

| Question | Answers (n=63) |
|---|---|
| 1. Are there any other adult individuals (age >=18 years old) who were referenced in this report as (likely) being affected with the genetic disease (e.g., there is mention of the variant being inherited from "parent with …", but no other details are provided and that parent is not participating in the study)? | Yes: 6.3%<br>No: 93.7% |
| 2. Was/were the adult(s) phenotyped in adulthood? | Always: 85.7%<br>Sometimes: 1.6%<br>Never: 12.7% |
| 3. Is there any description related to the adult's mental health status or quality of life? | Yes: 64.3%<br>No: 35.7% |
| 4. Is the level of functioning described using adult-appropriate (e.g., not "global developmental delay") language? | Yes: 64.3%<br>No: 35.7% |
| 5. Is highest level of educational achievement explicitly mentioned? | Always: 1.6%<br>Sometimes: 15.9%<br>Never: 82.5% |
| 6. Is their residence/living environment mentioned (e.g., group home, residential care, supported housing, lives independently)? | Always: 1.6%<br>Sometimes: 3.2%<br>Never: 95.2% |
| 7. Is their relationship status (past or present) mentioned? | Always: 1.6%<br>Sometimes: 1.6%<br>Never: 96.8% |
| 8. Was their reproductive history described? | Always: 3.2%<br>Sometimes: 1.6%<br>Never: 95.2% |
| 9. Is employment explicitly mentioned? | Always: 1.6%<br>Sometimes: 3.2%<br>Never: 95.2% |
| 10. Is their level of independence in activities of daily living mentioned (e.g., dressing, eating, toileting, chores/work, etc.)? | Always: 6.4%<br>Sometimes: 9.5%<br>Never: 84.1% |
| 11. Is driver license attainment mentioned? | Always: 0%<br>Sometimes: 0% |

| | |
|---|---|
| | Never: 100% |
| 12. Is their primary caregiver mentioned / implied? | Always: 11.1%<br>Sometimes: 4.8%<br>Never: 84.1% |

**Supplementary Table 7. Phenotype extraction form data for Domain VI for the initial reports describing individuals with seizures/epilepsy.**

| Question | Answers (n=85) |
|---|---|
| 1. Was epilepsy considered by the authors to be a major feature of the condition (e.g., epilepsy is mentioned in the title or abstract)? | Yes: 30.6%<br>No: 69.4% |
| 2. Is seizure type(s) mentioned (at least at one timepoint)? | Always: 40%<br>Sometimes: 42.4%<br>Never: 17.6% |
| 3. Is the epilepsy syndrome(s) mentioned (at least at one timepoint)? | Always: 3.5%<br>Sometimes: 14.1%<br>Never: 82.4% |
| 4. Is age of onset of seizures mentioned? | Always: 42.4%<br>Sometimes: 34.1%<br>Never: 23.5% |
| 5. Are EEG findings explicitly described? | Always: 22.4%<br>Sometimes: 43.5%<br>Never: 34.1% |
| 6. Are any individuals mentioned who had sudden unexpected death in epilepsy, or otherwise died from an epilepsy-related complication? | Yes: 5.4%<br>No: 94.6% |
| 7. Are current anti-seizure medications (or lack thereof) mentioned? | Always: 22.4%<br>Sometimes: 42.5%<br>Never: 34.1% |
| a. Is dosage mentioned? | Always: 0%<br>Sometimes: 8.5%<br>Never: 91.5% |
| b. Is there mention of anti-seizure treatments that were trialed and then discontinued for any participant? | Yes: 31.8%<br>No: 68.2% |
| c. Is this a variable that was clearly documented for all participants with seizures/epilepsy? | Yes: 33.3%<br>No: 66.7% |
| d. Are adverse drug events (or lack thereof) mentioned that relate to anti-seizure treatments, for any participant? | Yes: 4.7%<br>No: 95.3% |

| | |
|---|---|
| e. Is this a variable that was clearly documented for all participants with seizures/epilepsy? | Yes: 0%<br>No: 100% |
| 8. Is there explicit mention for any participant of dietary interventions for seizures (e.g., ketogenic diet)? | Yes: 16.5%<br>No: 83.5% |
| a. Is this a variable that was clearly documented for all participants with seizures/epilepsy? | Yes: 21.4%<br>No: 78.6% |
| 9. Is there explicit mention for any participant of surgical interventions for seizures (e.g., VNS insertion, epilepsy surgery)? | Yes: 7.1%<br>No: 92.9% |
| a. Is this a variable that was clearly documented for all participants with seizures/epilepsy? | Yes: 16.7%<br>No: 83.3% |

**Supplementary Table 8. Summary of the modified Delphi process used to formulate the PHELIX_General version 1.0 reporting checklist.**

| **Stage** | **Date(s)** | **Total number of items discussed and ranked** | **Engagement** |
|---|---|---|---|
| **1. Systematic review of the existing literature** | January 2022 – December 2022 | Not applicable | Not applicable |
| **2. Electronic surveys** | January 2023 – April 2023 | 50 | 9 of 11 |
| **3. Large group meeting** | May 18, 2023 | 35 | Live meeting attendance: 6<br><br>Feedback after asynchronous viewing: 1 |
| **4. Final review and approval of checklist items** | August 2023 – September 2023 | See Table 1 for details | All listed co-authors |

**Supplementary Table 9. Expanded PHELIX_General (version 1.0) with examples and notes.**

| Domain | Recommendation Category | Recommendations | Examples/Notes |
|---|---|---|---|
| **1. Development and Cognition** | Strongly recommended | i. Standard and specific diagnostic term(s) for cognitive or developmental issue(s)<br><br>ii. Level of cognitive functioning or degree of developmental delay<br><br>iii. Age of attaining major milestones<br><br>iv. Quantitative results from psychometric testing OR explicit acknowledgement that these results were not available | PMID: 30609410 Refer to case description of family 1 in the article and supplementary file.<br><br>PMID: 30487643 Refer to case descriptions for individuals 1 and 3 in the article. |
| | Optional, but encouraged | v. Narrative summary describing progression / change in cognitive or development issue(s) over time. | |
| **2. Behaviour and Neuropsyhicatric Conditions** | Strongly recommended | i. Standard and specific diagnostic term(s) for behavioural issues | PMID: 30487643 Refer to case description for individual 2 |
| | Optional, but encouraged | ii. Reported functional impact of behavioural/psychiatric condition<br>iii. Age at diagnosis and/or age at first concern for behavioural issues<br>iv. Impact of treatments / interventions, as reported by individuals, families, and/or clinicians | |

| | | | |
|---|---|---|---|
| **3. Other Medical Conditions** | Strongly recommended | i.Major medical conditions<br><br>ii. Presence or absence of issues in the following areas (if potentially associated with the condition under study):<br><br>- Visual acuity and field of vision<br>- Hearing ability<br>- Speech/communication styles<br>- Continence/toileting<br>- Ambulation | PMID: 28692176<br>Refer to case descriptions of all patients in the article that consistenly mention the absence of hearing and vision issues. |
| **4. Feeding Issues** | Strongly recommended | i. Functional impact of feeding issues<br><br>ii. Current feeding method (e.g., oral, gastrostomy tube) | PMID: 30401460<br>Refer to case report for individual 1 in the supplementary file. |
| | Optional, but encouraged | iii. Age at first concern for feeding issues<br><br>iv. Interventions and supports for feeding issues (e.g., feeding tube support) | |
| **5. Growth** | Strongly recommended | i. Birth growth measurements AND gestational age-corrected centiles<br><br>ii. Growth measurements (absolute values and z-scores) at two or more post-birth timepoints (where possible) | PMID: 28920961<br>Refer to patient report 1 in the supplementary file. |
| **6. Medication and Treatment History** | Optional, but encouraged | i. Details of efficacious treatment(s)<br><br>ii. Severe adverse events / reactions | PMID: 32220291<br>Refer to case report for individual 2 in the supplementary file that reports on medications trialed for seizures. |
| **7. Pain, Sleep, and Quality of Life** | Optional, but encouraged | i. Presence or absence of pain / neuroirritability<br><br>ii. Presence or absence of abnormal sleep patterns / sleep disturbance | PMID: 33632309<br><br>Refer to Table 6 as examples of |

| | | | |
|---|---|---|---|
| | | iii. Qualitative description of proxies for quality of life, via patient and/or caregiver report<br><br>iv. Direct assessment of quality of life using established measure(s), via patient and/or caregiver report | reporting sleeping disorders<br><br>PMID: 36740581 |
| **8. Indicators of adult functional outcome** | Strongly recommended | i. Age at which the adult was last seen/phenotyped<br><br>ii. Description of educational achievement<br><br>iii. Nature of any employment (past and/or present) | PMID: 25569435, 36729053 |
| | Optional, but encouraged | iv. Relationship status (past and/or present)<br><br>v. Reproductive history | |
| **9.Other** | Strongly recommended | i. Confirmation of informed consent to participate in the research study and to include the above phenotype information, for each participant<br><br>ii. Distinguish between "not assessed" and "assessed and not present," for every aspect of a phenotype described in the report and for each participant<br><br>iii. Description of how phenotyping was performed (e.g., direct assessment by study team member(s), review of medical records, information provided on testing requisition), for each participant<br><br>iv. Use of standard phenotype labels (e.g., HPO, ICD-11) to standardize reporting, for each participant.<br><br>v. [For deceased participants] Cause of death | PMID: 34284219 |

Abbreviations: HPO, Human Phenotype Ontology; ICD-11, International Classification of Diseases 11th Revision

**Supplementary Table 10:** Expanded PHELIX_Epilepsy (version 1.0) reporting guideline with examples.

| Domain | Recommendation Category | Recommended Information To Include In Epilepsy / Seizure Phenotype Reporting (If Applicable In Clinical Context) | Examples/Notes |
|---|---|---|---|
| **1. Epilepsy syndrome and severity** | Strongly recommended | i. Seizure type(s) (per ILAE)<br>ii. Epilepsy syndrome(s) (per ILAE)<br>iii. Findings that support the diagnosis of the epilepsy syndrome(s) (e.g., specific EEG findings)<br>iv. Seizure frequency at last clinical assessment<br>v. Qualitative description of overall epilepsy severity | PMID: 28942967<br>Refer to Table 1 and accompanying Results section |
| | Optional, but encouraged | vi. Number of seizures requiring hospitalization in specific timeframe (e.g., last year) | |
| **2. Pharmacological interventions** | Strongly recommended | i. Current and past medication name(s)<br>ii. Perceived impact on seizure control | PMID: 32220291<br>Refer to case report for individual 2 in the supplementary file that reports on medications trialed for seizures. |
| | Optional, but encouraged | iii. Dose<br>iv. Duration of treatment trial<br>v. Adverse effects/events due to the intervention | |
| **3. Non-pharmacological interventions** | Strongly Recommended | i. Intervention / procedure details (e.g., ketogenic diet, neurosurgery)<br>ii. Perceived impact on seizure control | PMID: 28942967<br>Refer to Table 1 and accompanying Results section |
| | Optional, but encouraged | iii. Adverse effects/events due to the intervention | |
| **4. Brain imaging and EEG findings** | Strongly recommended | i. Brain imaging findings (including age(s) at time of study) | PMID: 30323019 |

| | | | |
|---|---|---|---|
| | | ii. EEG findings (including age(s) at time of study) | Refer to reports on patients 1 and 2 in the article. |
| | Optional, but encouraged | iii. Clarification if brain imaging and/or EEG data were directly reviewed by members of the study team (versus only report details extracted from medical record) | |
| **5. Other** | Optional, but encouraged | i. Narrative summary of the progression of the individual's seizure(s)/epilepsy phenotype over time<br>ii. Narrative summary of the progression of the individual's non-epilepsy phenotype over time (e.g., see PHELIX_General guidelines) | |
